# Using the LIST model to Estimate the Effects of Contact Tracing on COVID-19 Endemic Equilibria in England and its Regions

**DOI:** 10.1101/2020.06.11.20128611

**Authors:** Rosalyn J. Moran, Alexander J. Billig, Maell Cullen, Adeel Razi, Jean Daunizeau, Rob Leech, Karl J. Friston

## Abstract

Governments across Europe are preparing for the emergence from lockdown, in phases, to prevent a resurgence in cases of COVID-19. Along with social distancing (SD) measures, contact tracing – find, track, trace and isolate (FTTI) policies are also being implemented. Here, we investigate FTTI policies in terms of their impact on the endemic equilibrium. We used a generative model – the dynamic causal ‘Location’, ‘Infection’, ‘Symptom’ and ‘Testing’ (LIST) model to identify testing, tracing, and quarantine requirements. We optimised LIST model parameters based on time series of daily reported cases and deaths of COVID-19 in England— and based upon reported cases in the nine regions of England and in all 150 upper tier local authorities. Using these optimised parameters, we forecasted infection rates and the impact of FTTI for each area—national, regional, and local. Predicting data from early June 2020, we find that under conditions of medium-term immunity, a ‘40%’ FTTI policy (or greater), could reach a distinct endemic equilibrium that produces a significantly lower death rate and a decrease in ICU occupancy. Considering regions of England in isolation, some regions could substantially reduce death rates with 20% efficacy. We characterise the accompanying endemic equilibria in terms of dynamical stability, observing bifurcation patterns whereby relatively small increases in FTTI efficacy result in stable states with reduced overall morbidity and mortality. These analyses suggest that FTTI will not only save lives, even if only partially effective, and could underwrite the stability of any endemic steady-state we manage to attain.

## Introduction

The efficacy of contact tracing programs within the context of widespread outbreaks of COVID-19 have recently been considered (Aleta A et al 2020, Ferretti et al 2020, Giordano et al 2020, Gurdasani & Ziauddeen 2020, Hellewell et al 2020, Keeling et al 2020), with differing conclusions that depend upon the form of the models. Models that include social distancing measures—along with quarantine of infected contacts—show that a ‘find’, ‘track’, ‘trace’ and ‘isolate’ (FTTI) policy can ameliorate infection rates (Giordano et al 2020, Kretzschmar et al 2020). Investigating endemic equilibria is important when considering FTTI policies. For example, a steady state of a certain number of cases per day could be achieved under social distancing policies alone. An FTTI policy could simply delay this steady state, by allowing susceptible individuals to re-enter a community epidemic. However, another alternative is that an FTTI policy can actually improve death rates at steady state (i.e., endemic equilibrium). Here we aim to test the hypothesis that an improved endemic equilibrium can be engendered through FTTI.

Quantifying the public health resources needed for testing and contact tracing is a necessary prerequisite for implementing a successful FTTI policy. Though more difficult to predict, likely compliance should also be considered. In 2009, for example, in the context of a newly introduced (H1N1) virus in Melbourne, Australia, compliance was 85% of those quarantined (McVernon et al 2011).

In order to accommodate our evolving scientific understanding of SARS-CoV-2, including, for example seroprevalence (Vespignani et al 2020) or immunity status (Kissler et al 2020), planning frameworks require adaptable, real-time predictive capacity. The dynamic causal LIST (location, infection, symptom, and testing) model offers this capacity. It can also be applied as an inverse model—whereby historical data of recorded cases and deaths can be used to optimise model parameters, providing a mechanistic summary of how these data were generated, including specific testing regimes. The inferred parameters can then be used to forecast the near-future epidemic landscape. This rests on the fact that the LIST model is a generative model (Friston et al 2020a). Previously we have employed LIST to examine local outbreak dynamics (Friston et al 2020a), the susceptible populations involved in the outbreaks (Moran et al 2020) and the likelihood of a second wave, in the context of resistant individuals (Friston et al 2020b, Friston et al 2020c).

Forecasting from the LIST model could be applied to support adaptive planning in two ways. Firstly, as an indicator system for prevalence of infection in a community, LIST, unlike regression frameworks can indicate the causes of R_0_, the reproductive number (or R_t_ – the effective reproduction number) in real-time, with daily estimates. Importantly the LIST model includes self-organising features—in particular, a mechanism whereby social distancing is coupled to infection rates in the community. Having these metrics available for regional communities could, in principle, enable a local adaptive response.

In what follows, we report the results of simulated FTTI-based policies, incorporating social distancing. We rehearse the established LIST model and show how it can recapitulate the dynamics of the epidemic at national, regional, and sub-regional levels. We simulate several levels of ‘success’ of a tracing policy, investigating the impact of identifying and isolating asymptomatic virus carriers. We sought to investigate the endemic equilibria under various levels of contact tracing efficacy. We found that epidemiological (population) dynamics can subtend distinct equilibria depending on the FTTI parameter. Crucially, we discovered that certain FTTI policies produce an equilibrium that includes significantly lower death rates and lower ICU occupancy. Technically speaking, we found that there was a critical level of FTTI efficacy that, in the rhetoric of dynamical systems theory, plays the role of a *bifurcation parameter*, which determines whether we end up with a manageable endemic equilibrium or a second epidemiological steady-state, with much greater mortality rates. In other words the level of effective FTTI reaches a critical value at which point mortality rates drop dramatically after small improvements in FTTI.

## Methods

### Data

Data from Public Health England (PHE) (https://coronavirus.data.gov.uk/) were used to inform model parameters and thereby infer the latent causes of these data. Time series of reported cases of COVID-19 for the nation of England were recorded from Jan 30^th^, 2020, (1^st^ confirmed case). We analysed data up to May 18^th^, 2020. Reports of COVID-19 confirmed deaths ‘*Deaths of people who have had a positive test result confirmed by a Public Health or NHS laboratory’* accompanied the case report data for England with the first death recorded on the 9^th^ of March, 2020. The total cases reported in England by PHE up to this date at the time of analysis was 146,273. The total deaths reported at the time of analysis was 31,008. Population sizes for the nation, regions and upper tier local authorities were obtained from the 2018 census reports (https://www.ons.gov/peoplepopulationandcommunity).

Regional time series comprised confirmed cases only, from the nine regions of England: the North West, the North East, Yorkshire and the Humber, the West Midlands, the East Midlands, the East of England, London, the South West and the South East. Death records were not divided by regions or local authority. Confirmed case data from Upper Tier Local Authorities (UTLA) for all regions were also analysed for these dates. These comprised:

#### 23 Local Authorities of the North West

Blackburn with Darwen, Blackpool, Bolton, Bury, Cheshire East, Cheshire West and Chester, Cumbria, Halton, Knowsley, Lancashire, Liverpool, Manchester, Oldham, Rochdale, Salford, Sefton, St. Helens, Stockport, Tameside, Trafford, Warrington, Wigan, Wirral,

#### 15 Local Authorities of Yorkshire and the Humber

Barnsley, Bradford, Calderdale, Doncaster, East Riding of Yorkshire, Kingston upon Hull, Kirklees, Leeds, North East Lincolnshire, North Lincolnshire, North Yorkshire, Rotherham, Sheffield, Wakefield, York

#### 12 Local Authorities of the North East

County Durham, Darlington, Gateshead, Hartlepool, Middlesbrough, Newcastle upon Tyne, North Tyneside, Northumberland, Redcar and Cleveland, South Tyneside, Stockton-on-Tees, Sunderland,

#### 14 Local Authorities of the West Midlands

Birmingham, Coventry, Dudley, County of Herefordshire, Sandwell, Shropshire, Solihull, Staffordshire, Stoke-on-Trent, Telford and Wrekin, Walsall, Warwickshire, Wolverhampton, Worcestershire

#### 9 Local Authorities of the East Midlands

Derby, Derbyshire, Leicester, Leicestershire, Lincolnshire, Northamptonshire, Nottingham, Nottinghamshire, Rutland

#### 11 Local Authorities of the East of England

Bedford, Cambridgeshire, Central Bedfordshire, Essex, Hertfordshire, Luton, Norfolk, Peterborough, Southend-on-Sea, Suffolk, Thurrock

#### 33 Local Authorities of London

Barking and Dagenham, Barnet, Bexley, Brent, Bromley, Camden, City of London, Croydon, Ealing, Enfield, Greenwich, Hackney, Hammersmith and Fulham, Haringey, Harrow, Havering, Hillingdon, Hounslow, Islington, Kensington and Chelsea, Kingston upon Thames, Lambeth, Lewisham, Merton, Newham, Redbridge, Richmond upon Thames, Southwark, Sutton, Tower Hamlets, Waltham Forest, Wandsworth, Westminster

#### 14 Local Authorities of the South West

Bath and North East Somerset, ‘Bournemouth, Christchurch and Poole’, City of Bristol, Cornwall and Isles of Scilly, Devon, Dorset, Gloucestershire, North Somerset, Plymouth, Somerset, South Gloucestershire, Swindon, Torbay, Wiltshire

#### And 19 Local Authorities of the South East

Bracknell Forest, Brighton and Hove, Buckinghamshire, East Sussex, Hampshire, Isle of Wight, Kent, Medway, Milton Keynes, Oxfordshire, Portsmouth, Reading, Slough, Southampton, Surrey, West Berkshire, West Sussex, Windsor and Maidenhead, Wokingham.

### LIST Model

The LIST model comprises four factors describing the state of an individual in the population. These states describe four potential Locations: ‘At Home’, ‘At Work’, ‘In the ICU’ or ‘Deceased’, five potential Infection states: ‘Susceptible’, ‘Infected’, ‘Infectious’, ‘Immune’ and ‘Resistant’, four potential Symptom states: ‘Asymptomatic’, ‘Symptomatic’, ‘Acute Respiratory Disease (ARDS)’ and ‘Deceased’ and four Testing states: ‘Untested’, ‘Waiting’, ‘Positive Test Result, and ‘Negative Test Result’. Thus, the combinatorics allow for an individual to occupy 320 distinct states e.g. at home, infected, with symptoms, while waiting for a test.

Probabilistic state transitions occur within factors determining, for example, the probability of developing ARDS once symptomatic. These sorts of probabilities and other rate parameters—for example the time to recover from infection given mild symptoms—are set to *a priori* levels consistent with the literature (Chen et al 2020, Wang et al 2020a, Wang et al 2020c, Weiss & Murdoch 2020). Parameters are subsequently optimised for each time series or time series pair, for example, the probability of transferring from ICU is set to the national average *a priori* but will be optimised for each locale to produce ICU admission rates that best explain the local data. Dynamics, i.e., time dependent changes in cases and deaths, unfold via tensor operations on these transition matrices. The *a priori* probabilities of state transitions, are given in Appendix 1. Importantly some transitions within a factor are dependent on other factors—i.e. second order dependencies—for example where the probability of fatality from ARDS is dependent on location, whether ‘at home’ (which could include care homes), or in an ICU.

### Parameter Optimisation

In order to create an optimal LIST model for the nation, for each region and for each local authority, we first optimised the parameters based on the national data for England, which comprises two time series of cases per day and deaths per day. Then we used these optimised parameters as priors on the regional and local authority data. Parameter priors comprise a multivariate Gaussian density described by its mean and covariance. A table of these values is provided in Appendix 1. Both national mean and covariance matrices were applied as priors to the regional and local authority data; since for regional and local authority data, only case reports were available for parameter optimization. This is appropriate given that parameters can describe the generative mechanisms of case and death outputs or only case outputs – but will be better constrained *a priori* using both. The optimization scheme employs a standard (Variational Laplace) Bayesian scheme that performs a gradient descent on the variational free energy (a.k.a. evidence lower bound) of the model (Friston et al 2007). This cost function provides constraints on fits (precision weighted errors on predicted data) while minimising complexity, i.e., the divergence of posterior estimates from prior values.

We fit all 160 models (150 local authorities, 9 regions and 1 nation), using the time series from Jan 30^th^ to May 18^th^, 2020. Figure 3 shows the predicted responses against the empirical time series. Crucially, we use the posterior parameters (that do not change with time) to produce forecasts of latent or hidden states (that do change with time) into early June. From the predicted states of May 18th (including location, infection and symptom states) we then generated forecasts for a range of FTTI parameters accompanied by adaptive (self-organising) social distancing.

### FTTI in the LIST Model

To model a find track and trace policy, the LIST model conditions the probability of being tested on whether or not you are infected – and can augment the relative testing probability if you are infected and asymptomatic – akin to testing a successfully-traced contact. Then the total probability of being tested is a weighted average of two probabilities, such that

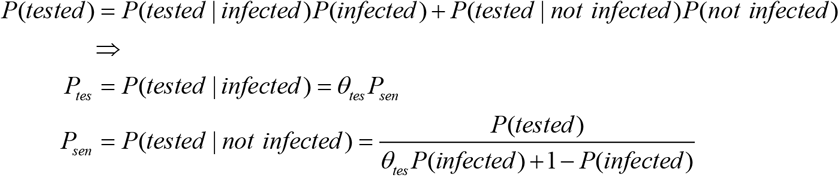

where the probability of being tested has three components:

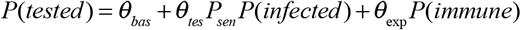

where the first component *θ*_*bas*_ is a *baseline* probability of receiving a test, the second component parameterises an increase in testing rates as the prevalence of infection in the community increases and the third term is a *sustained* response (using the level of immunity as a proxy for a gently declining function of the cumulative number of affected people).

To model targeted testing of ‘contacts’ through a contact tracing scheme, we parameterise an enhanced probability of testing if, and only if, you are infected and asymptomatic. Noting that the proportions tested below pertain not to all those contacted but to those contacted, infected, and asymptomatic. By adding this probability to the probability of being tested when infected, we supplement general screening with an FTTI parameter as follows:

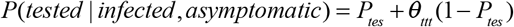

In the absence of any targeted testing, efficacy will be zero. A priori, the efficacy was set to very low levels of one in 10,000 people, per day – to model the data to May 18^th^ when tracking and tracing was not commonplace. We adjust *θ*_*ttt*_, the efficacy of testing, from 0 to 0.05, 0.3, 0.4, 0.5 and 0.6 to simulate the long-term horizon or endemic equilibria that would ensue from identifying different proportions of asymptomatic individuals through contact tracing.

Given this model of FTTI and its efficacy, we used the remaining parameters available from fitting the model to data from each nation, region, and local authority to project forward from May 18^th^ to measure the impact of these various FTTI policies. We furthermore adjust the immunity parameter to assess the impact of various levels of immunity conferred by previous infection – assessing long (32 month), medium (16 month), short term (3 month) and no (0 month) immunity.

Importantly, these policies subsume a propensity to leave the house (*Pount*) based upon the levels of infection in the community. This means we are effectively testing combined policies of FTTI under adaptive social distancing.

## Results

To quantify the impact of FTTI policies, we used the optimised parameters from the original LIST models. Specifically, we harvested a snapshot of the predicted states from May 18^th^ 2020, obtaining probabilities for each of the states in the Location (L), Infection (I) Symptom (S) and Testing (T) factors of each model (national, regional and local authority). From here we projected outcomes for 36 months under different FTTI policies: we varied the efficacy *θ*_*ttt*_ parameter from 0 (effectively no targeted testing) to 5%, 20%, 40%, 50% and 60%.

Using the national model, we first examined high immunity levels after infection (32-month immunity). With this, we observed that for policies of 20% efficacy and greater, a distinct endemic equilibrium emerged with substantially lower infections, deaths, and ICU occupancy (Figure 1). For a more modest 16-month immunity, a 40% efficacy was required. For this 16-month immunity assumption, the model predicted that relative to the 0%, 5% or 20% policy, a reduction of 15 to 25 thousand deaths would be obtained (by May 18^th^ 2022). Similarly, ICU day rates would fall significantly (Figure 1). For lower length immunity following infection these distinct equilibria were not evidenced (3-month and 0-month immunity durations, Figure 1).

**Figure 1.**
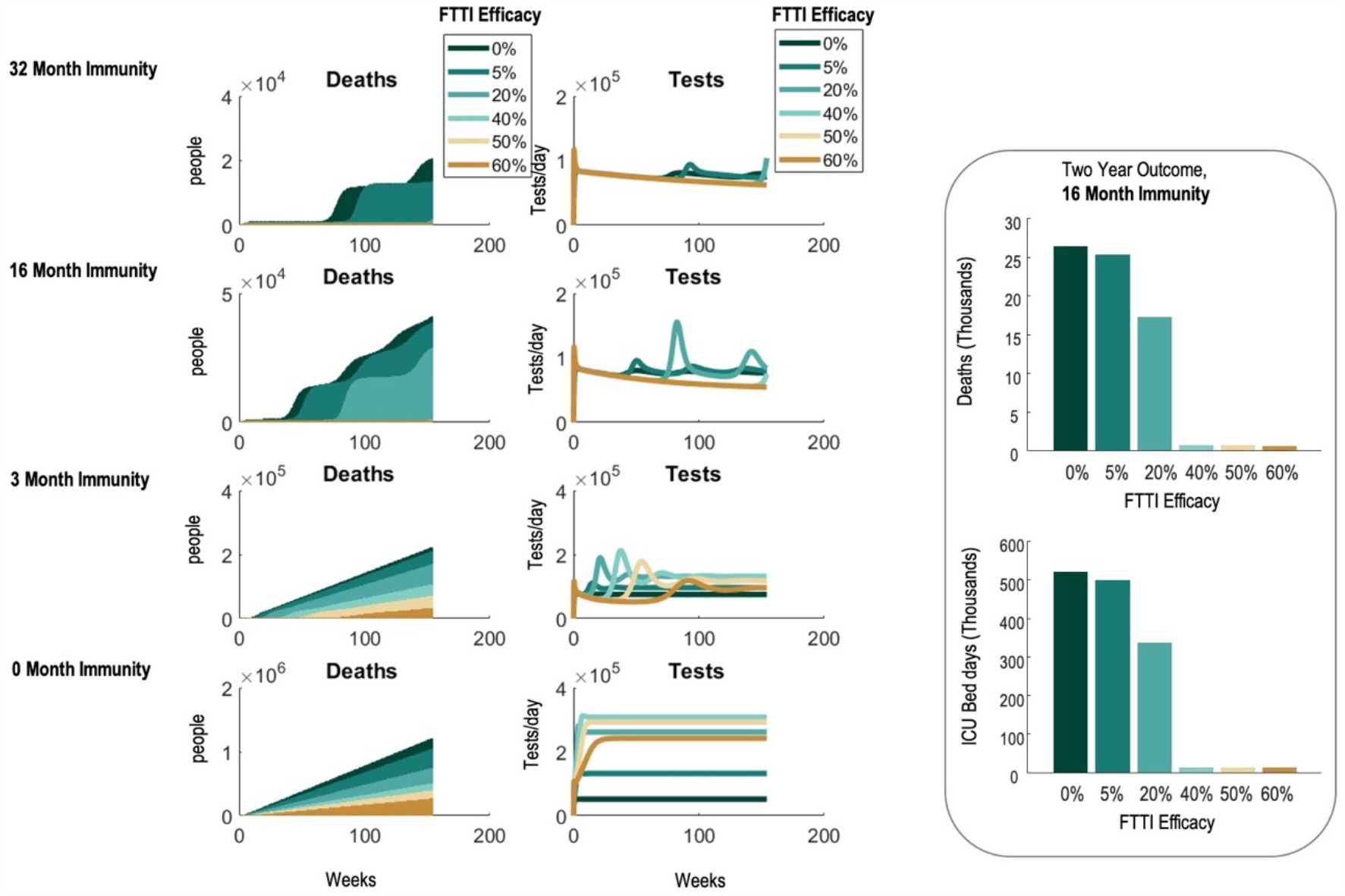
FTTI Policies under various levels of immunity. Projected outcomes for England after May 18^th^ 2020 under various quarantine policies. Six policies are illustrated with 0%, 5%, 20%, 40%, 50% and 60% efficacy. We observe distinct endemic equilibria, where a bifurcation is evident when changing the efficacy parameter under the conditions of longer-term and medium-term immunity. The left panels show the cumulative death toll under the six policies. The middle panels show the associated daily testing rates. The right panel examines death rates and ICU bed days that would accumulate until May 18^th^ 2022 under the various policies assuming 16 months immunity.

For the 16-month and 0-month immunity model we explored the dynamics further to ensure that they have distinct stationary points (i.e., endemic fixed points). Using simulations over long periods of time, we aimed to investigate the steady state behaviour of the models – rather than for predictive purposes. We show that both FTTI and no quarantine policies reach a steady state with a stability analysis, revealing greater dynamical stability with FTTI (Figure 2A). We characterised these fixed points by converting the transfer functions to the z-domain and examining the associated pole-zero plots (appropriate for high dimensional systems). From our perspective, we are interested in the poles (Figure 2B, respectively). A negative real part of the pole reflects the stability of the steady state—or the rate of convergence to steady-state. It can be seen that with 16 months of immunity the 50% efficacy has a more stable steady-state, relative to the no quarantine, 0% efficacy policy (Figure 2B).

**Figure 2.**
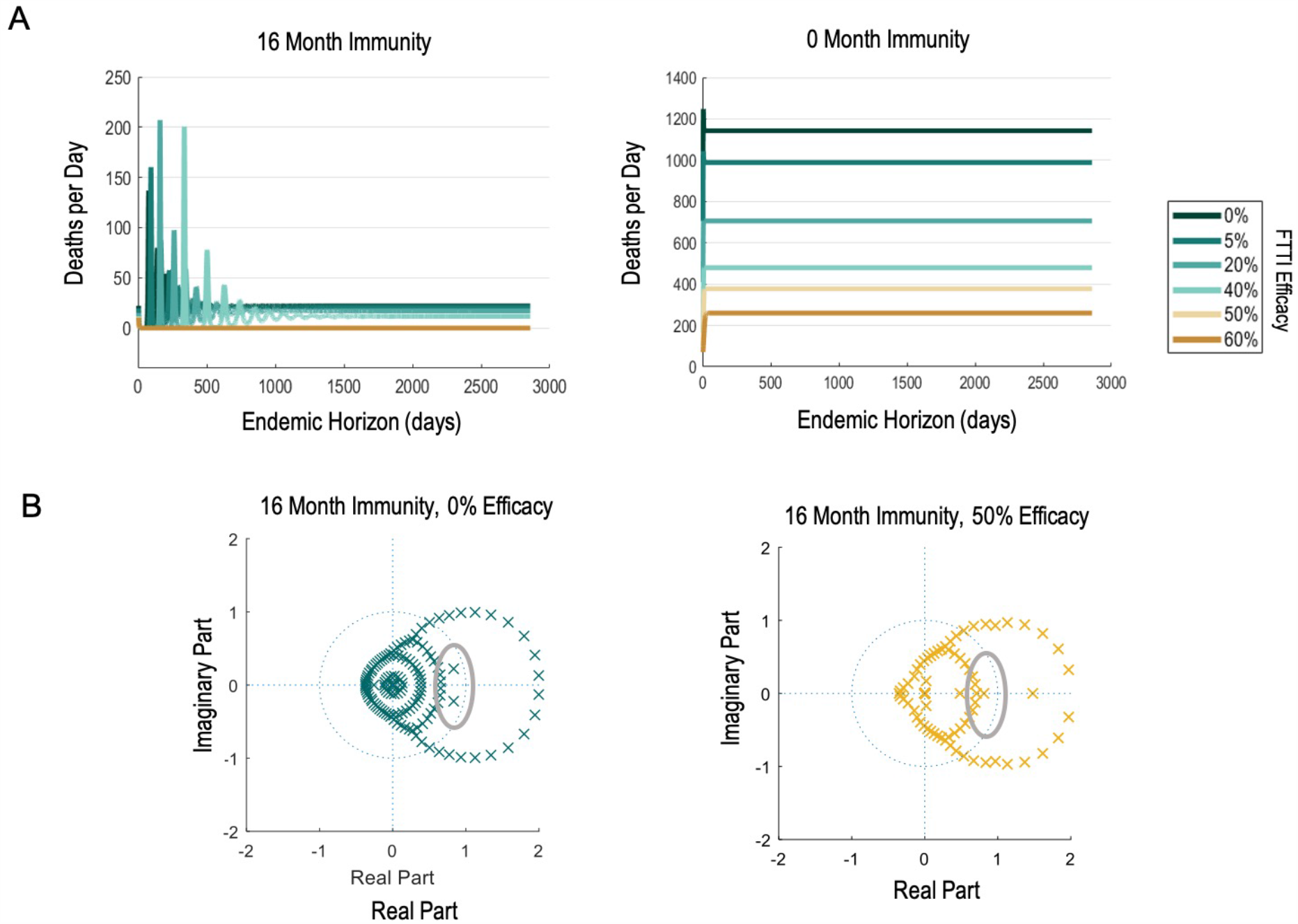
Endemic Equilibria. We examined the steady state behaviour in terms of dynamical stability for medium term and no immunity scenarios. Upper panels: Extended time course for the system under varying levels of FTTI efficacy exhibit distinct steady states. Lower panel: pole-plots for the 16-month immunity case two models reveals poles closer to the unit circle for 0% efficacy relative to 50% efficacy – indicating increased instability at the lower FTTI efficacy level. Poles found outside of the unit circle typically induce instability but here are accompanied by zeros (not shown) which engender stability. We highlight the poles within the unit circle in grey circles to show that though both stable regimes, the poles in green at 0% efficacy are closer to the unit circle and with higher frequency, whereas the pole in yellow at 50% efficacy is farther away from instability and with zero frequency (along the x axis).

**Figure 3.**
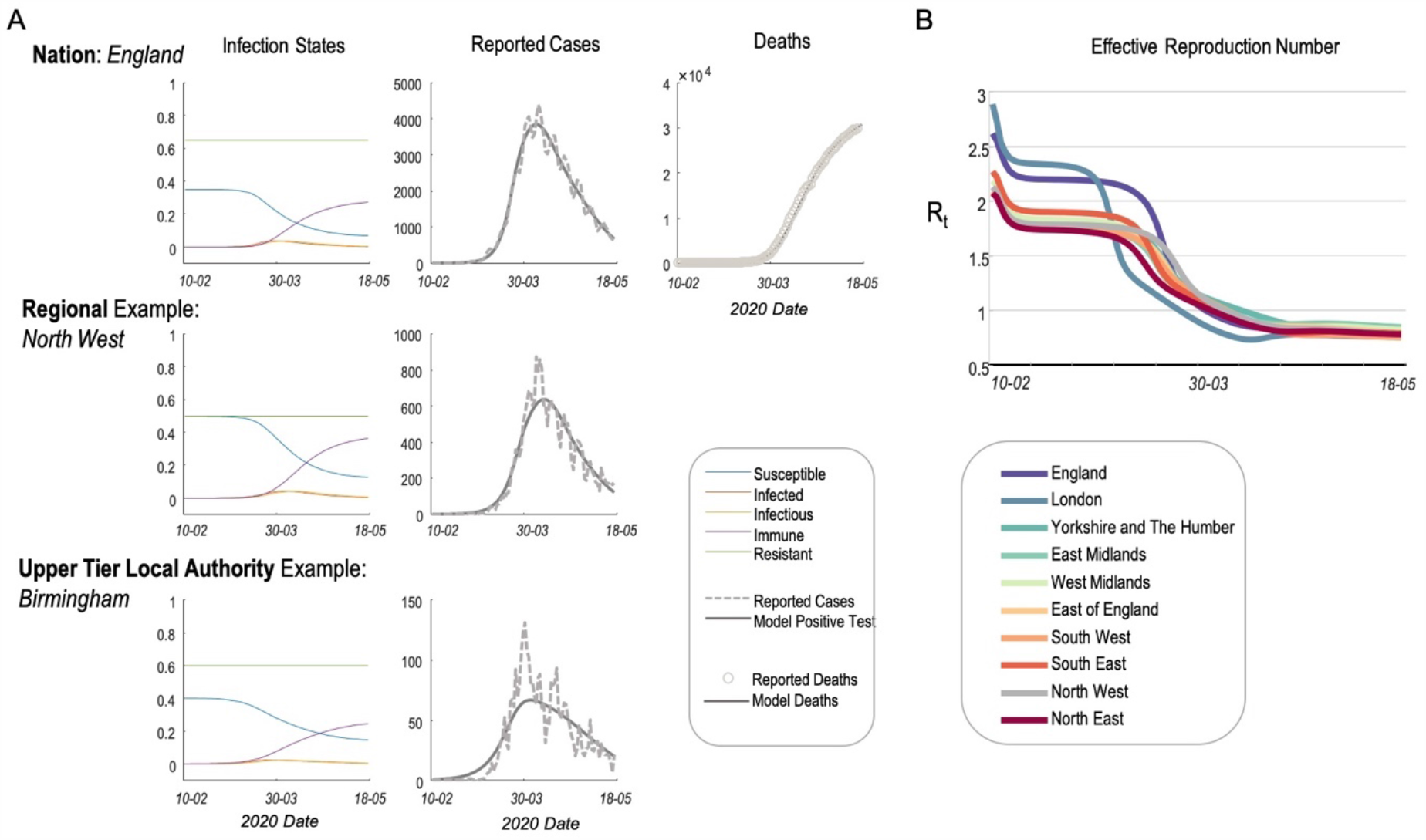
LIST model optimised for England, regions of England and Local Authorities. Left Panel: Modelled latent states underlying the national cases as well as the regional example of the North West and the UTLA example of Birmingham. Mid Left Panels: Empirical data are shown as dashed lines and model predictions with full lines. Empirical reports up to May 18^th^, 2020 were analysed. Deaths were available for the national model (upper panel). Right: Region specific effective reproduction rates (R_t_).

To characterise regional variations in the spread of the virus – and the effect of FTT I – we sought to establish whether the model could reproduce the dynamics of recently reported outbreaks at regional and upper-tier local authority level. Using the original LIST model we optimised the parameters based on reported cases and deaths (England) and based upon the reported cases for each region (the North West, the North East, Yorkshire and the Humber, the West Midlands, the East Midlands, the East of England, London, the South West and the South East) and local authority (UTLA) within those regions. Prior values of parameters were set as previously reported (Friston et al 2020a) and are provided in Appendix A. We find that the models accurately recapitulate the dynamics of these time series, capturing outbreaks (from Jan 30^th^ to May 18^th^) in the regions and upper tier local authority data in a similar way to the national data (Figure 3).

It is important to note that the models account for a proportion of the population that are not susceptible to infection in the current outbreak but could partake in future outbreaks. These ‘resistant’ proportions (discussed below) were set a priori to 0.25 and were found to be 0.64 for the national model and on average 0.42 ± 0.06 (*mean* ± *std*) once optimised to the nine regional datasets.

To account for social distancing, the model features an infection level-dependent probability of leaving home. For near-future SD policies that adapt to individual area levels of infection, we examined the instantaneous reproduction numbers (R_t_) to highlight their time-sensitivity. We find that the model predicts larger initial reproduction rates for London than for England as a whole. We observe differing levels of R_t_ for different regions where, for example, London exhibits a higher R_t_ early on in the outbreak relative to Yorkshire, but Yorkshire takes ∼2 weeks longer for the R_t_ to fall below 1 (Figure 3).

Given our regional models, we then investigated whether regions exhibited similar bifurcation patterns in terms of FTTI efficacy to each other and to the Nation (Figure 4). We found that considering each region as a closed system resulted in a bifurcation to low death rates at lower efficacy rates than when using a national model. Specifically, a policy of 40% would reduce deaths significantly in the North West, North East, East Midlands, Yorkshire and the Humber, The West Midlands, the South East, the East of England and in the South West. However, a policy of 50% efficacy would be required for London in the long term (“Regional Bifurcations” in Figure 4).

**Figure 4.**
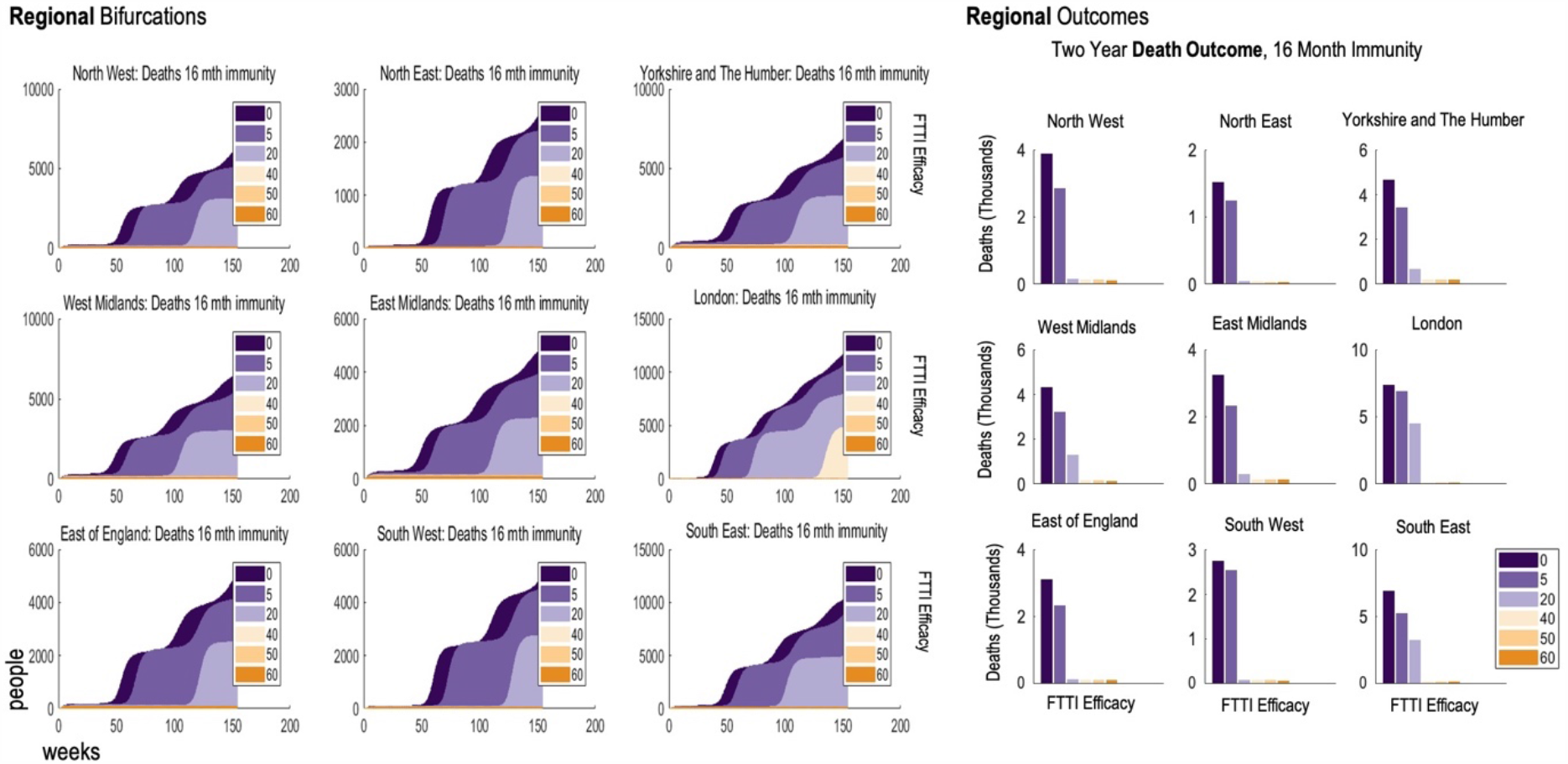
Regional FTTI Policies. Projected outcomes for the regions of England after May 18^th^ 2020, under various FTTI policies and assuming 16 months of immunity. Six policies are illustrated with 0%, 5%, 20%, 40%, 50% and 60% efficacy. Again, we observe that low policies delay deaths over a three-year period (left panel) but policies of 40% and higher reduced deaths. In London a 50% policy is required for significant death reduction. The right panels show the death rates that would accumulate until May 18^th^ 2022 under the various policies assuming 16 months immunity.

Given these regional results, we used the regional testing capacity above the bifurcation level in an attempt to inform ‘shoe leather’ epidemiology in each local authority. We examined the predicted testing rates (using the initial testing levels at day 1 after the implementation of the policy, rather than those at steady state, which reduce from the initial testing rates – see Figure 1). Using the UTLA models for each region (Figure 5) but assuming that the closed system was at a regional level – i.e. requiring the testing percentages shown in Figure 4, we show the local authorities sorted into predicted testing levels.

**Figure 5.**
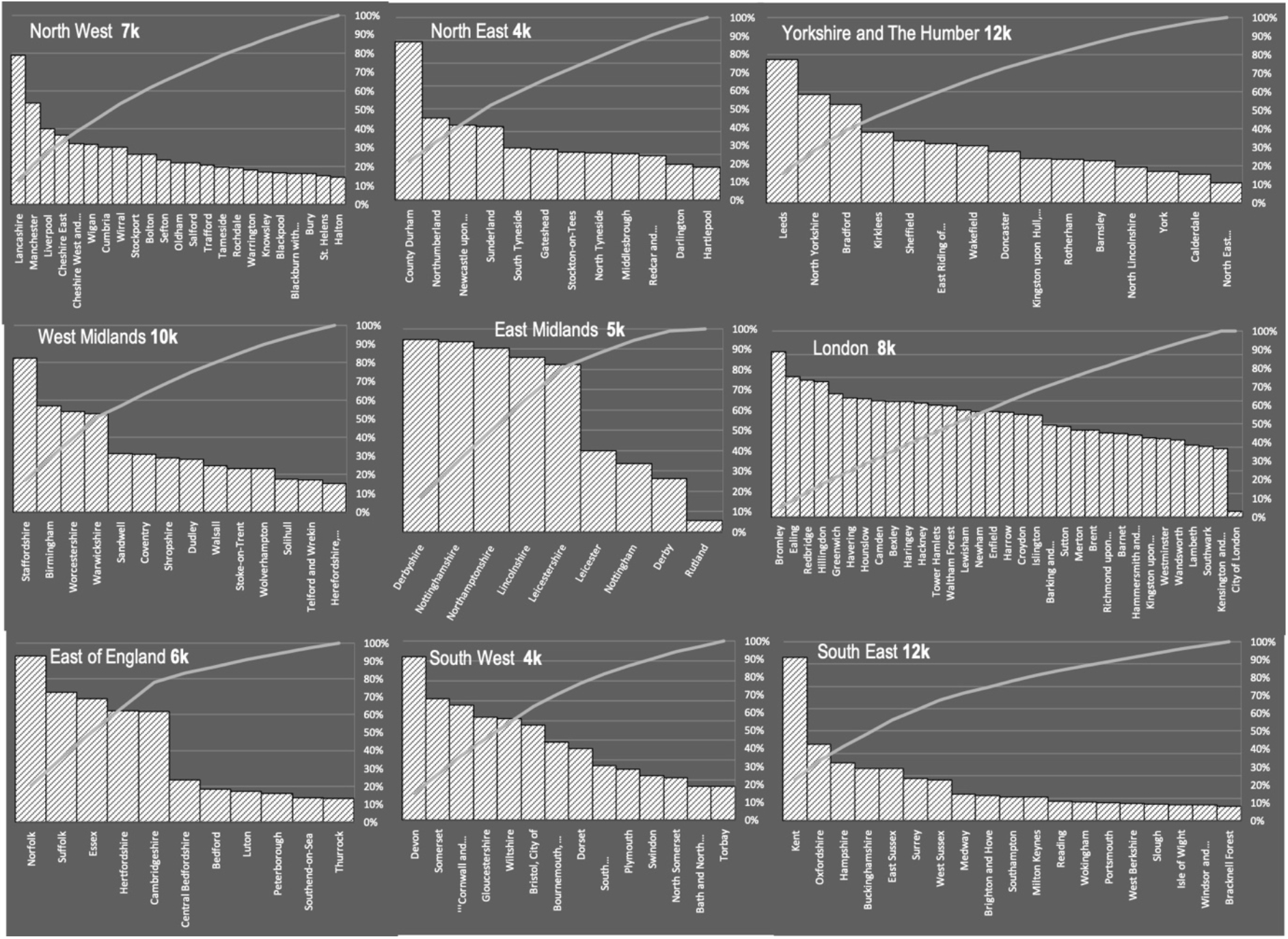
Predicted Daily Testing Requirements for Upper Tier Local Authorities at Early Phases of FTTI policy. We assume the corresponding testing levels from each region (Figure 4) are a cumulative count of testing with the UTLAs, where the regional testing amount is 100% (y axis). We used the UTLA models to apportion the testing levels required in each region. We illustrate ‘day 1’ testing requirements which would decrease to lower daily testing rates at steady state (see Figure 1 for an exemplar testing trajectory).

Interestingly the UTLAs with maximum predicted testing requirements (Figure 5) were not necessarily those where the most cases have been identified in the past. For example, in London the highest testing requirements are predicted to be in Bromley (Figure 5), while Croydon has had the highest number of cases.

## Discussion

We have used a dynamic causal (LIST) model to forecast the impact of a combined FTTI and social distancing policy for COVID-19 infection and death prevention. Our findings speak to the possibility of a feasible FTTI policy that would significantly ameliorate COVID-19 related deaths and ICU occupancy in England. Importantly we identified non-linearities that place hard constraints on the success of an FTTI policy—with a bifurcation from a low death rate endemic equilibrium to a higher (though delayed) death rate at an efficacy below 40% under assumptions of medium-term immunity following infection. Identifying these bifurcation points is useful for simultaneously minimising the level of FTTI policy and social distancing while minimising disease burden and death.

We applied a LIST model used to investigate trajectories of outbreaks in the UK (Friston et al 2020a), the potential for second waves (Friston et al 2020b, Moran et al 2020) and the spread of COVID-19 across America (Friston et al 2020b). The latter study demonstrated two important features for both strategic planning and near-future policies related to decision making around COVID-19. Firstly, it exposed different sensitivities in social distancing. This is operationalized by placing the probability of leaving a small number of contacts (‘at home’) for a larger number of contacts (‘at work’) as a function of the levels of infection prevalence in the community. This self-organising policy is included in our model. Secondly the study revealed that, in the US, local, state-wide policies would result in improved outcomes (in terms of deaths and cases) as opposed to a federal policy, where the sensitivities are linked to levels of infection across multiple regions. In light of these findings we produced model-based estimates for local authorities, rather than for national level statistics. The interplay of epidemic parameters with endogenous behavioural change has also been observed in the case dynamics of Hubei and other affected Chinese provinces, with interpretations that large fractions of these populations remain at risk (Maier & Brockmann 2020, Tian et al 2020).

In terms of supporting a community warning system, the instantaneous reproduction rate R_t_, is an important and widely understood metric. Here, we show that the LIST model can generate R_t_ in real-time, using the latent infection rate. Furthermore, it can be forecast from these models (Figure 3). In a recent investigation of the impact of social distancing and lockdown measures, robust R_t_ estimates were derived though an hierarchical Bayesian Regression using known covariates (regressors) corresponding to different stages of lockdown, with interventions shown to have profound effects on disease transmission (Flaxman et al 2020). The LIST model may be a useful adjunct to these approaches by incorporating FTTI parameters. More generally, our results highlight the non-intuitive non-linearities present in these epidemic dynamics; for example, by doubling the efficacy of quarantining, more than double the number of lives may be saved.

A potentially useful perspective on this modelling is afforded by dynamical systems theory. We found that there was a critical level of FTTI that plays the role of a *bifurcation parameter*, which determines the endemic equilibrium. In this simplified and idealised model, the endemic equilibria are generally fixed points. However, the stability of these endemic equilibria appears to be sensitive to interventions like FTTI. This is potentially relevant if one considers the epidemiological response of the population to perturbations (for example, the arrival of infected people from another country). In short, not only may FTTI save lives, it may underwrite the stability of any endemic equilibrium we manage to attain.

### Limitations

There are several limitations to the analysis. First, the data used to optimise the models would ideally consider death rates, rates of hospitalisations and rates of ICU occupancy as well as symptomatic and asymptomatic infections for all regions. Since the only data publicly available for regions and UTLAs were case reports – the model is fallible to incorrect or incomplete reporting as well as omitted data. Second, our results are predicated on a generative model of the epidemic course of COVID-19 (Daunizeau et al 2020) which should be refined based on biological assays of the community. For example, the overall rates of infection within UK’s UTLAs (Winter & Hegde 2020, Yong et al 2020) particularly given the low seroprevalence in global epicentres (To et al 2020). Most crucially, immunity after infection (where here, we assume a relatively long period of immunity) has yet to be established (Kissler et al 2020). Furthermore, the models account for a proportion of the population that are not susceptible to infection in the current outbreak, i.e. only a subset of the population are in the susceptible compartment at the beginning of the outbreak. Possible causes of ‘resistance’ as estimated by the model include: geographical effects, i.e. not involved in current outbreaks through lockdown measures and social sequestering from the mixing population (Flaxman et al 2020), innate host factors e.g., under expression of the angiotensin-converting enzyme 2 (ACE2) gene in children (Bunyavanich et al 2020), endogenous immunity through cross immunity with historical betacoronavirus infections (Grifoni et al 2020), reduced transmission of asymptomatic carriers (Davies et al), or overdispersion of transmission c.f., super-spreaders and cluster events (Shim et al 2020). Thus, our results pertain to outbreaks involving a subset of a population with far less than 100% of the population open to infection.

Overall, we observe that a large initial FTTI effort in the emergence from an epidemic cycle may preclude a protracted and more severe infection in the proceeding months – perhaps sustaining a ‘new normal’ that is closer to the old normal.

## Data Availability

All data used are publicly available and provided in the data availability link

https://zenodo.org/badge/latestdoi/271566597

## Software Reproducibility

To reproduce the analysis and results presented here, code, datasets and metadata can be found at:

https://zenodo.org/badge/latestdoi/271566597

## Appendix 1

Prior Parameters of the LIST Model. Prior means are for scale parameters *θ* exp(*ϑ*)

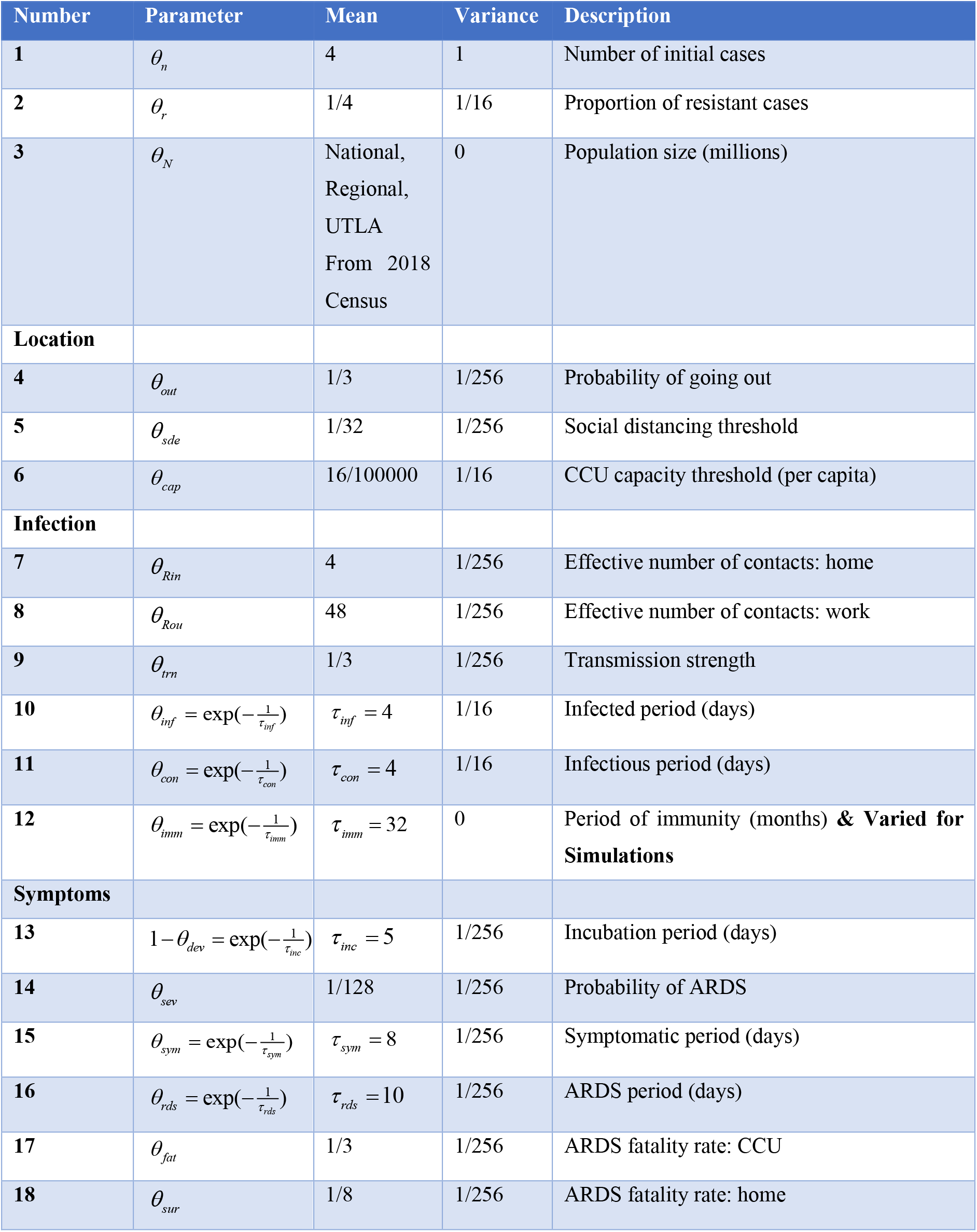
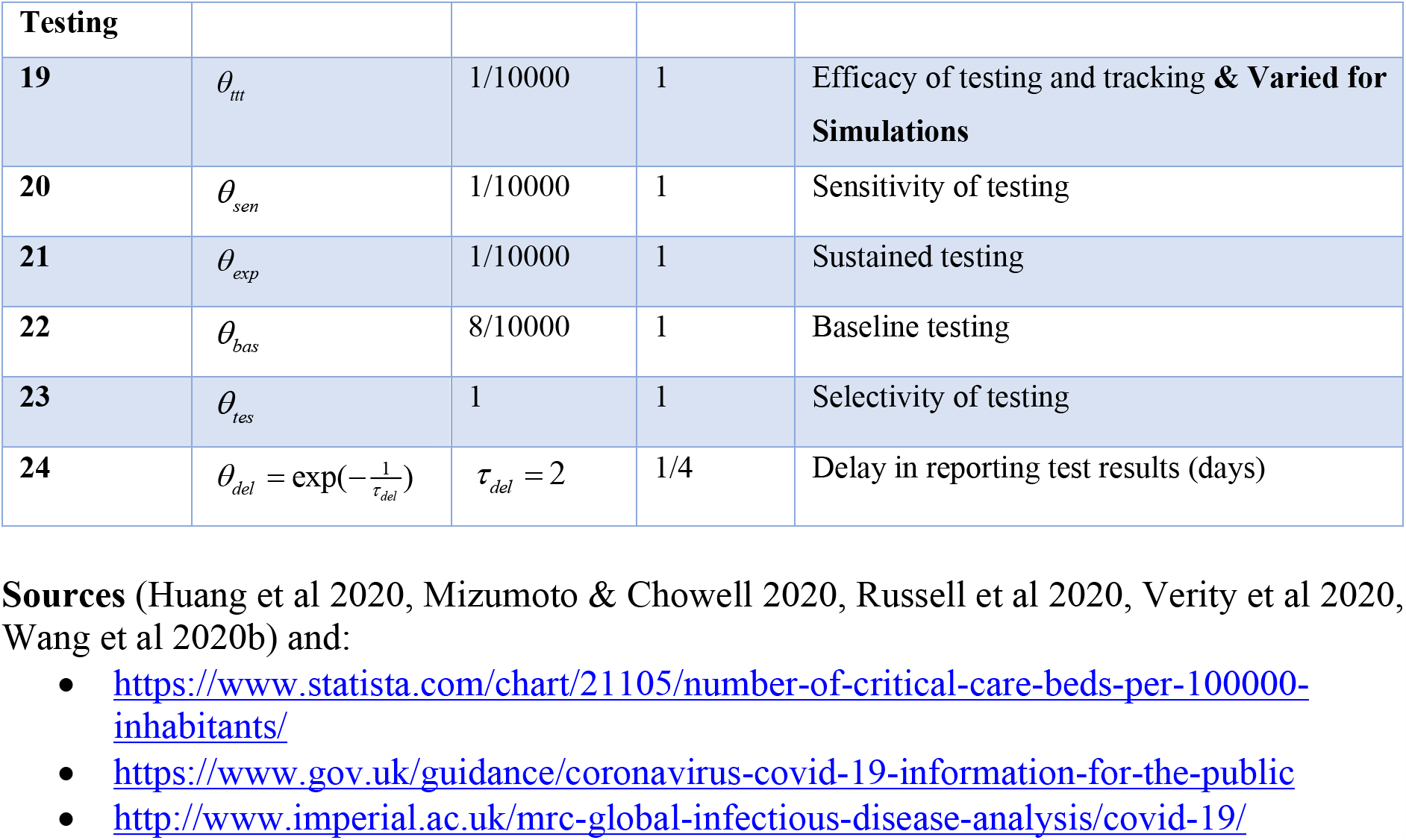

## Notes

### Competing Interest Statement

The authors have declared no competing interest.

### Funding Statement

No external funding

### Author Declarations

Exemption, public data analysis

## References

1. Aleta A, Martin-Corral C, Pastore Y Pionttia A, Jelli M, Litvinova M, et al. 2020. Modeling the impact of social distancing, testing, contact tracing and household quarantine on second-wave scenarios of the Covid-19 epidemic. In cidid

2. Bunyavanich S, Do A, Vicencio A. 2020. Nasal gene expression of angiotensin-converting enzyme 2 in children and adults. JAMA

3. Chen N, Zhou M, Dong X, Qu J, Gong F, et al. 2020. Epidemiological and clinical characteristics of 99 cases of 2019 novel coronavirus pneumonia in Wuhan, China: a descriptive study. The Lancet 395: 507–13

4. Daunizeau J, Moran RJ, Mattout J, Friston K. 2020. On the reliability of model-based predictions in the context of the current COVID epidemic event: impact of outbreak peak phase and data paucity. medRxiv

5. Davies NG, Kucharski AJ, Eggo RM, Gimma A, Edmunds WJ, et al. Effects of non-pharmaceutical interventions on COVID-19 cases, deaths, and demand for hospital services in the UK: a modelling study. The Lancet Public Health

6. Ferretti L, Wymant C, Kendall M, Zhao L, Nurtay A, et al. 2020. Quantifying SARS-CoV-2 transmission suggests epidemic control with digital contact tracing. Science

7. Flaxman S, Mishra S, Gandy A, Unwin HJT, Mellan TA, et al. 2020. Estimating the effects of non-pharmaceutical interventions on COVID-19 in Europe. Nature

8. Friston K, Mattout J, Trujillo-Barreto N, Ashburner J, Penny W. 2007. Variational free energy and the Laplace approximation. Neuroimage 34: 220–34

9. Friston KJ, Parr T, Zeidman P, Razi A, Flandin G, et al. 2020a. Dynamic causal modelling of COVID-19 [version 1; peer review: awaiting peer review]. Wellcome Open Res 5:89

10. Friston KJ, Parr T, Zeidman P, Razi A, Flandin G, et al. 2020b. Second waves, social distancing, and the spread of COVID-19 across America. arXiv preprint 2004.13017

11. Friston KJ, Parr T, Zeidman P, Razi A, Flandin G, et al. 2020c. Second waves, social distancing, and the spread of COVID-19 across America. arXiv preprint 2004.13017

12. Giordano G, Blanchini F, Bruno R, Colaneri P, Di Filippo A, et al. 2020. Modelling the COVID-19 epidemic and implementation of population–wide interventions in Italy. Nature Medicine: 1–6

13. Grifoni A, Weiskopf D, Ramirez SI, Mateus J, Dan JM, et al. 2020. Targets of T cell responses to SARS-CoV-2 coronavirus in humans with COVID-19 disease and unexposed individuals. Cell

14. Gurdasani D, Ziauddeen H. 2020. On the fallibility of simulation models in informing pandemic responses. The Lancet Global Health

15. Hellewell J, Abbott S, Gimma A, Bosse NI, Jarvis CI, et al. 2020. Feasibility of controlling COVID- 19 outbreaks by isolation of cases and contacts. The Lancet Global Health

16. Huang CL, Wang YM, Li XW, Ren LL, Zhao JP, et al. 2020. Clinical features of patients infected with 2019 novel coronavirus in Wuhan, China. Lancet (London, England) 395: 497–506

17. Keeling MJ, Hollingsworth TD, Read JM. 2020. The Efficacy of Contact Tracing for the Containment of the 2019 Novel Coronavirus (COVID-19). medRxiv

18. Kissler SM, Tedijanto C, Goldstein E, Grad YH, Lipsitch M. 2020. Projecting the transmission dynamics of SARS-CoV-2 through the postpandemic period. Science

19. Kretzschmar M, Rozhnova G, van Boven M. 2020. Isolation and contact tracing can tip the scale to containment of COVID-19 in populations with social distancing. Available at SSRN 3562458

20. Maier BF, Brockmann D. 2020. Effective containment explains subexponential growth in recent confirmed COVID-19 cases in China. Science

21. McVernon J, Mason K, Petrony S, Nathan P, LaMontagne AD, et al. 2011. Recommendations for and compliance with social restrictions during implementation of school closures in the early phase of the influenza A (H1N1) 2009 outbreak in Melbourne, Australia. BMC Infectious Diseases 11: 257

22. Mizumoto K, Chowell G. 2020. Estimating Risk for Death from 2019 Novel Coronavirus Disease, China, January-February 2020. Emerging infectious diseases 26

23. Moran RJ, Fagerholm ED, Cullen M, Daunizeau J, Richardson MP, et al. 2020. Estimating required ‘lockdown’cycles before immunity to SARS-CoV-2: model-based analyses of susceptible population sizes,‘S0’, in seven European countries, including the UK and Ireland. Wellcome Open Research 5: 85

24. Russell TW, Hellewell J, Jarvis CI, van Zandvoort K, Abbott S, et al. 2020. Estimating the infection and case fatality ratio for coronavirus disease (COVID-19) using age-adjusted data from the outbreak on the Diamond Princess cruise ship, February 2020. Euro surveillance : bulletin Europeen sur les maladies transmissibles = European communicable disease bulletin 25

25. Shim E, Tariq A, Choi W, Lee Y, Chowell G. 2020. Transmission potential and severity of COVID-19 in South Korea. International Journal of Infectious Diseases

26. Tian H, Liu Y, Li Y, Wu C-H, Chen B, et al. 2020. An investigation of transmission control measures during the first 50 days of the COVID-19 epidemic in China. Science 368: 638–42

27. To KK-W, Cheng VC-C, Cai J-P, Chan K-H, Chen L-L, et al. 2020. Seroprevalence of SARS-CoV-2 in Hong Kong and in residents evacuated from Hubei province, China: a multicohort study. The Lancet Microbe

28. Verity R, Okell LC, Dorigatti I, Winskill P, Whittaker C, et al. 2020. Estimates of the severity of coronavirus disease 2019: a model–based analysis. The Lancet. Infectious diseases

29. Vespignani A, Tian H, Dye C, Lloyd-Smith JO, Eggo RM, et al. 2020. Modelling COVID-19. Nature Reviews Physics: 1-3

30. Wang D, Hu B, Hu C, Zhu F, Liu X, et al. 2020a. Clinical characteristics of 138 hospitalized patients with 2019 novel coronavirus–infected pneumonia in Wuhan, China. Jama 323: 1061–69

31. Wang D, Hu B, Hu C, Zhu F, Liu X, et al. 2020b. Clinical Characteristics of 138 Hospitalized Patients With 2019 Novel Coronavirus–Infected Pneumonia in Wuhan, China. JAMA 323: 1061–69

32. Wang H, Wang Z, Dong Y, Chang R, Xu C, et al. 2020c. Phase-adjusted estimation of the number of coronavirus disease 2019 cases in Wuhan, China. Cell Discovery 6: 1–8

33. Weiss P, Murdoch DR. 2020. Clinical course and mortality risk of severe COVID-19. The Lancet 395: 1014–15

34. Winter AK, Hegde ST. 2020. The important role of serology for COVID-19 control. The Lancet Infectious Diseases

35. Yong SEF, Anderson DE, Wei WE, Pang J, Chia WN, et al. 2020. Connecting clusters of COVID- 19: an epidemiological and serological investigation. The Lancet Infectious Diseases

